# Heterogeneous metabolomic aging across the same age and prediction of health outcome

**DOI:** 10.1101/2024.04.22.24306156

**Authors:** Xueqing Jia, Jiayao Fan, Xucheng Wu, Xingqi Cao, Lina Ma, Zeinab Abdelrahman, Daniele Bizzarri, Erik B van den Akker, P. Eline Slagboom, Joris Deelen, Dan Zhou, Zuyun Liu

## Abstract

Existing metabolomic clocks exhibit deficiencies in capturing the heterogeneous aging rates among individuals with the same chronological age. Yet, the modifiable and non-modifiable factors in metabolomic aging have not been systematically studied. Here, we leveraged metabolomic profiles of 239,291 UK Biobank participants for 10-year all-cause mortality prediction to generate and validate a new aging measure--MetaboAgeMort. The MetaboAgeMort showed significant associations with all-cause mortality, cause-specific mortality, and diverse incident diseases. Adding MetaboAgeMort to conventional risk factors model improved the predictive ability of 10-year mortality. We identified 99 modifiable factors for MetaboAgeMort, where 16 factors representing pulmonary function, body composition, socioeconomic status, dietary quality, smoking status, alcohol intake, and disease status showed quantitatively stronger associations. The genetic analyses revealed 99 genomic risk loci and 271 genes associated with MetaboAgeMort. Our study illuminates heterogeneous metabolomic aging across the same age, which provides avenues for developing anti-aging therapies and personalized interventions.

## Introduction

Aging is the greatest common risk factor for most chronic diseases^1^. The rate at which individuals age is heterogeneous, engendering variations in the susceptibility and progression of diseases and mortality^2^. As the global demographic shifts towards an aging population, the ability to measure aging, identify individuals who age faster, and understand the factors that contribute to differential rates of aging are of utmost importance. These findings have significant implications for the development of targeted preventive programs and interventions. These efforts may alleviate the socioeconomic and healthcare burden of age-related diseases, thus promoting healthy aging and longevity.

Metabolomics offers a novel avenue for assessing the biological processes that underlie aging^3^. To date, multiple studies have attempted to elucidate how metabolomic profiles in various tissues (e.g., blood, urine, and cerebrospinal fluid) interact with aging, and a few metabolomic clocks have been proposed to measure biological aging^4–7^. The majority of these clocks are generated based on correlations between metabolomic profiles and chronological age; while chronological age is considered an imperfect surrogate for building aging measures as it does not fully capture the heterogeneity of individual aging rates^8^. In contrast, metabolomic aging measures based on health-related surrogate indicators (e.g., time to death) may better reflect an individual’s health status and reveal intrinsic biological aging mechanisms^9^. Previous studies have utilized targeted or untargeted mass spectrometry and nuclear magnetic resonance (NMR) techniques to construct multivariable metabolite scores of all-cause mortality^10–12^. While these metabolite scores exhibit exceptional predictive accuracy even over conventional risk factor models, their applicability in risk stratification, especially across individuals of the same chronological age, remains constrained by their reliance on scaled biomarker values created independently for each cohort^12^. Metabolomic profiles manifest pronounced responsiveness to the confluence of endogenous genetic regulation and exogenous environmental exposures in each individual cohort^13^. Thus, developing aging measures that can be calculated based on concentration units derived from individual-level data, may have more applicability in both clinical setting and research on the biology of aging.

Although certain behaviors (e.g., dietary quality) have displayed anti-aging properties in human and animal models^14,15^, their impact on metabolomic aging remains uncertain. Furthermore, the complex relationship between aging and a constellation of diverse factors (e.g., local environmental factors and socioeconomic status [SES]) has emerged as a burgeoning focus in the field of aging research. A thorough investigation of these modifiable factors in metabolomic aging could reveal new strategies for preventive interventions targeting the aging process. Moreover, prior investigations have indicated that aging measures may capture distinct aging domains influenced by varying genetic determinants^16^. However, the underlying mechanisms and pathways of metabolomic aging have not been elucidated, thereby restricting the identification of potential therapeutic targets^17^.

In this study, we leveraged large-scale metabolomic data from the UK Biobank (UKB), a prospective cohort study of over 500,000 participants. We first developed a novel aging measure, MetaboAgeMort, using all-cause mortality as a surrogate^18^. Next, we evaluated its applicability by examining its association with aging-related outcomes (i.e., morbidity and mortality), and comparing its performance to conventional risk factors and several previously trained metabolomic clocks. Finally, we systematically identified the modifiable factors and genetic determinants for MetaboAgeMort (Fig. 1).

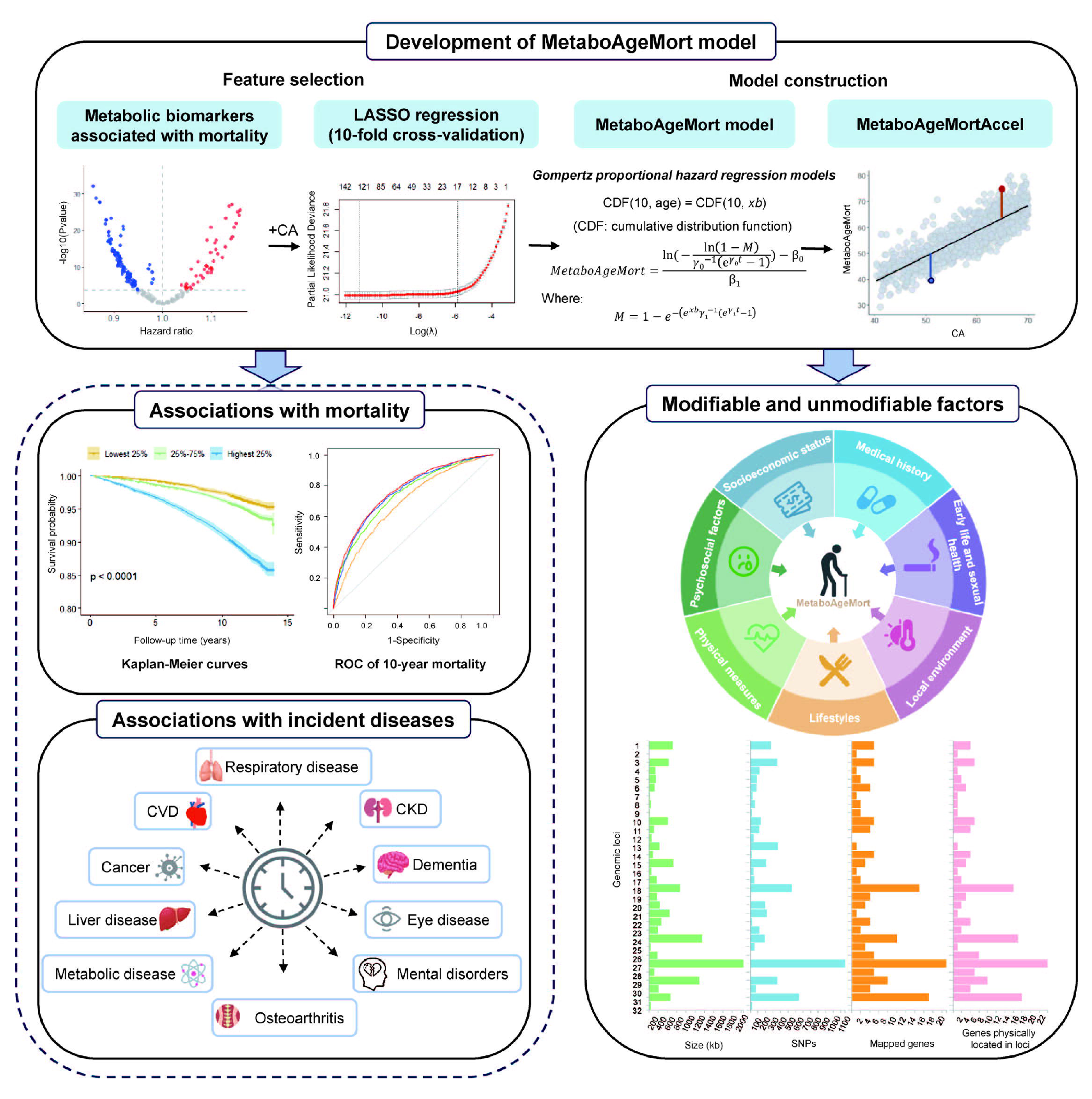

## Results

### Population characteristics

As shown in Supplementary Fig. 1, 239,291 participants with complete data on plasma metabolomics and covariates at baseline were included. These participants had a median age of 58.3 years (interquartile range [IQR]: 50.6, 63.7), and the majority were female (53.0%), and white ethnicity (95.6%). To develop the MetaboAgeMort model, the 239,291 participants were randomly split into a training (n = 167,506) and a testing set (n = 71,785), with a 7 to 3 ratio. No significant differences were observed in the sociodemographic characteristics of participants between the training set and the testing set. Detailed characteristics of the total participants and by datasets are presented in Supplementary Table 1.

### Development of MetaboAgeMort and MetaboAgeMort Acceleration (MetaboAgeMortAccel)

During a median follow-up of 13.9 years, we documented 20,447 deaths among 239,291 participants. After adjustment for potential confounders and accounting for multiple testing, a total of 185 metabolic biomarkers, encompassing amino acids, glycolysis-related metabolites, ketone bodies, fatty acids, lipids, and lipoprotein subclasses, demonstrated significant correlations with all-cause mortality (*P* < 0.05/249) (Supplementary Table 2).

To further select variables for inclusion in the MetaboAgeMort model, we applied a Cox regression model with least absolute shrinkage and selection operator (LASSO) penalization--where the hazard of all-cause mortality was regressed on the 185 metabolic biomarkers and chronological age--in the training set. Finally, chronological age and 35 metabolic biomarkers, including average diameter for very low-density lipoprotein (VLDL) particles, linoleic acid, ratio of omega-3 fatty acids to total fatty acids, ratio of monounsaturated fatty acids to total fatty acids, ratio of linoleic acid to total fatty acids, alanine, histidine, leucine, valine, phenylalanine, tyrosine, glucose, pyruvate, citrate, 3-hydroxybutyrate, acetate, acetoacetate, acetone, creatinine, albumin, glycoprotein acetyls, triglycerides in very large VLDL, free cholesterol in very large high-density lipoprotein (HDL), total lipids in small HDL, cholesteryl esters in small HDL, triglycerides to total lipids ratio in large VLDL, phospholipids to total lipids ratio in very small VLDL, phospholipids to total lipids ratio in intermediate density lipoprotein (IDL), cholesteryl esters to total lipids ratio in IDL, triglycerides to total lipids ratio in large low-density lipoprotein (LDL), triglycerides to total lipids ratio in medium LDL, phospholipids to total lipids ratio in small LDL, free cholesterol to total lipids ratio in small LDL, cholesteryl esters to total lipids ratio in very large HDL, free Cholesterol to total lipids ratio in small HDL, were selected (Supplementary Table 3).

Next, MetaboAgeMort was developed using the methods previously proposed by Levine et al. in the training set^18^. For more information about the MetaboAgeMort estimator, refer to Supplementary Table 4. A profiling of the MetaboAgeMort performance was carried out in the testing set (n = 71,785). MetaboAgeMort ranged from 27.82 to 104.06 years, with a mean and median value of 55.81 (standard deviation [SD] = 9.18) and 56.10 (IQR: 48.99, 62.50) years. As shown in Fig. 2a, MetaboAgeMort was highly correlated with chronological age across all participants (r = 0.85) and within each sex subgroup (female: r = 0.85; male: r = 0.86).

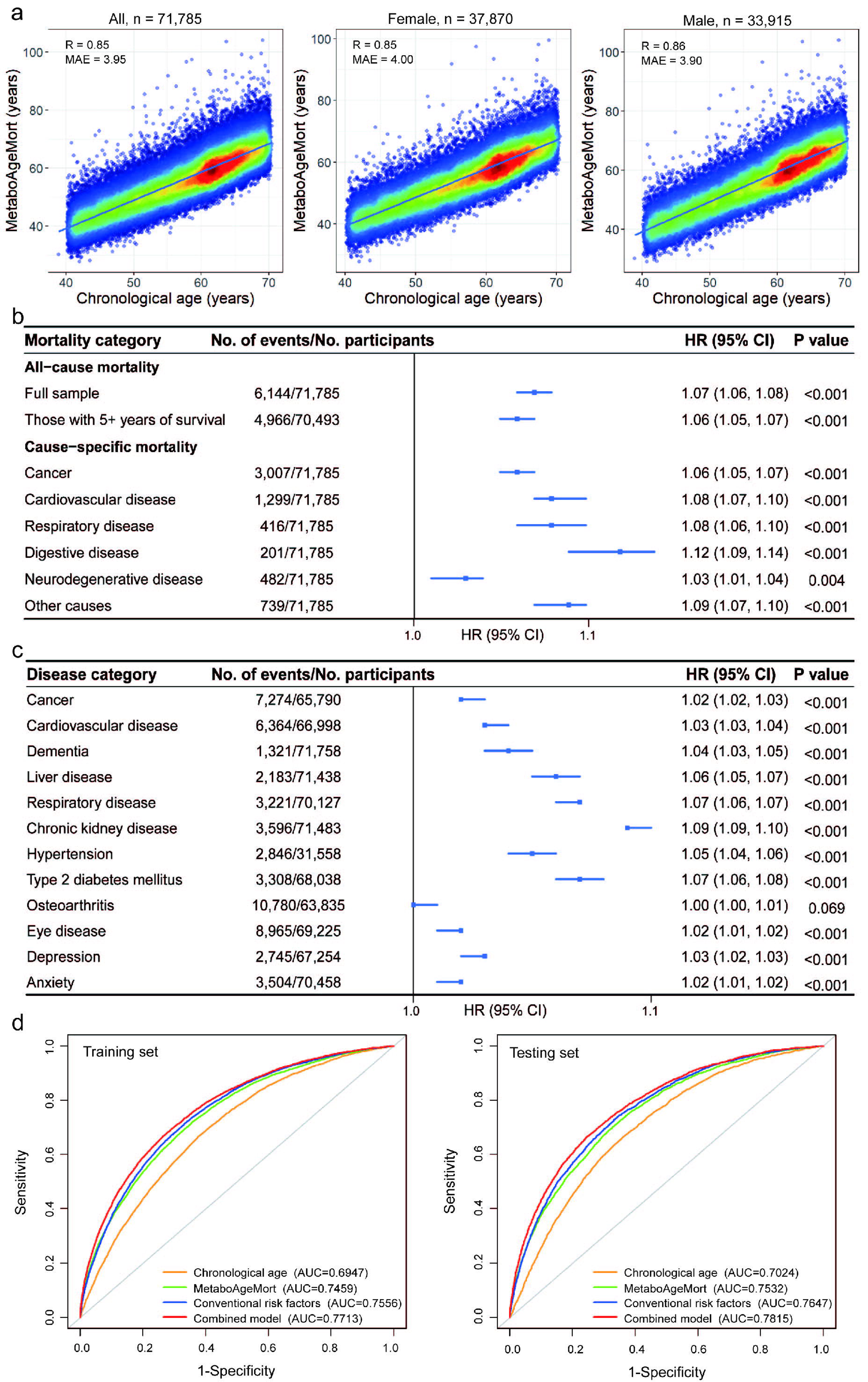

We also calculated a metric, MetaboAgeMortAccel, following the methods by Liu et al.^19^ MetaboAgeMortAccel represents the divergence of MetaboAgeMort from chronological age (i.e., whether a person appears younger [values < 0] or older [values > 0] than expected, based on his/her chronological age). The MetaboAgeMortAccel displayed a range of −16.43 to 41.29 years, with a mean and median value of 0 (SD = 4.78) and −0.41 (IQR: −3.29, 2.79) years in the testing set.

### Association of MetaboAgeMort with mortality

The associations of MetaboAgeMort with all-cause and cause-specific mortality in the testing set are demonstrated in Fig 2b. After adjustment for potential confounders, each additional year in MetaboAgeMort corresponded to an 7% rise in the risk of all-cause mortality (Hazard Ratio [HR]: 1.07, 95% confidence interval [CI]: 1.06, 1.08) (Supplementary Table 5). Our finding remains consistent when: (1) stratified by chronological age, sex, ethnicity, education level, smoking status, alcohol intake frequency, regular exercise, healthy diet, and body mass index (BMI) category (Supplementary Table 6), (2) excluding participants who died within five years of follow-up (Fig. 2b and Supplementary Table 5), and (3) restricting the sample to diseases-free participants (Supplementary Table 7). In addition, MetaboAgeMort was significantly positively associated with cause-specific mortality, including cancer (HR: 1.06, 95% CI: 1.05, 1.07), cardiovascular disease (CVD, HR: 1.08, 95% CI: 1.07, 1.10), respiratory disease (HR: 1.08, 95% CI: 1.06, 1.10), digestive disease (HR: 1.12, 95% CI: 1.09, 1.14), neurodegenerative disease (HR: 1.03, 95% CI: 1.01, 1.04), and other causes (HR: 1.09, 95% CI: 1.07, 1.10) (Fig. 2b and Supplementary Table 5).

The Kaplan-Meier survival curves indicated that individuals in the highest quartile group (Q4) of MetaboAgeMortAccel had significantly increased risks of all-cause and cause-specific mortality when compared to those in the lowest quartile (Q1) (Fig. 3a and Supplementary Table 8). The all-cause mortality rates of the highest quartile group (Q4) were found to be comparable, or in certain instances higher, than those of the lowest quartile group (Q1), despite the latter being 10 years older chronologically (Supplementary Fig. 4).

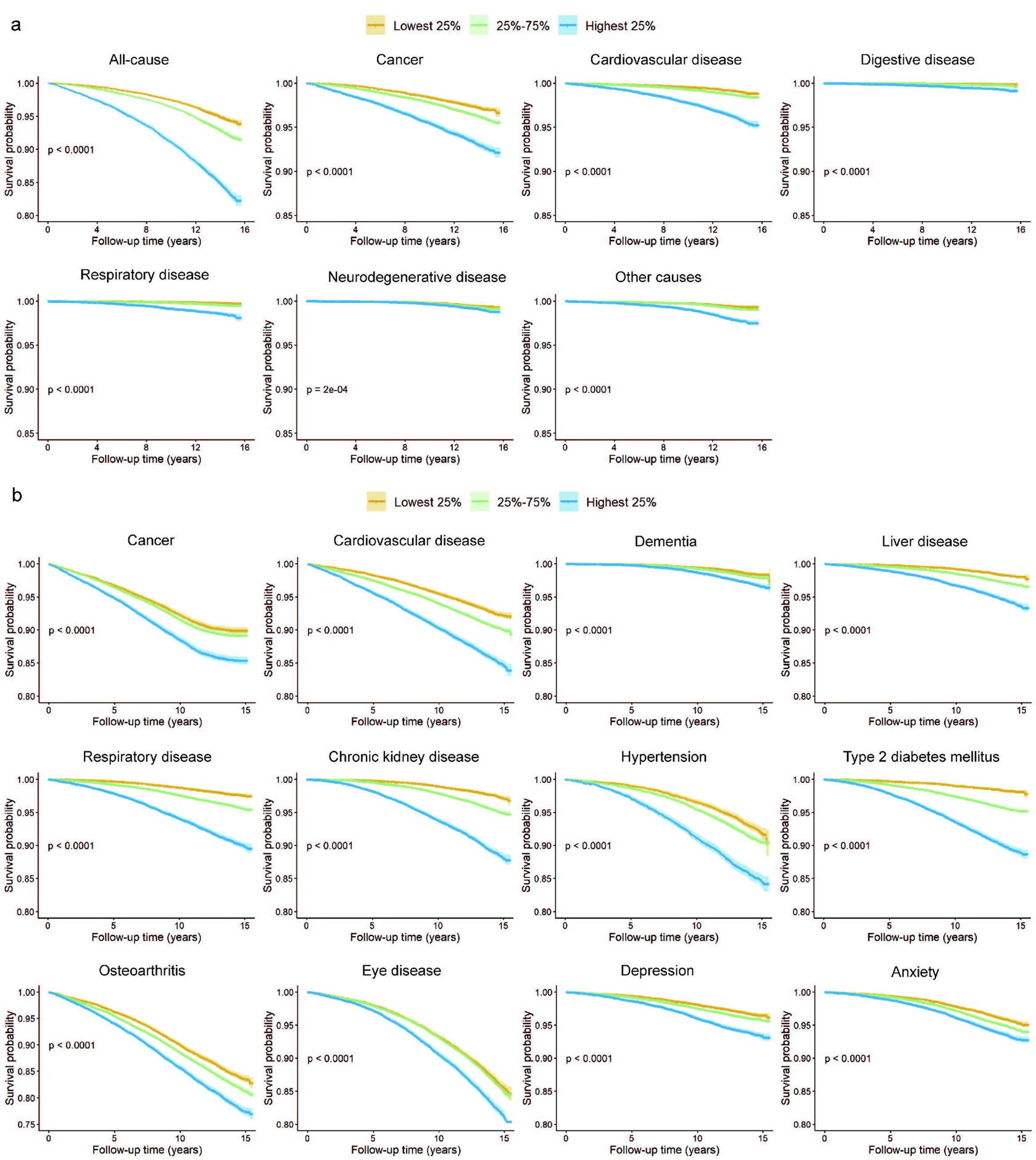

### Associations of MetaboAgeMort with diseases incidence

The associations of MetaboAgeMort with the risk of multiple diseases in the testing set are depicted in Fig. 2c. After adjustment for potential confounders, each 1-year increment in MetaboAgeMort was significantly associated with higher risks of cancer (HR: 1.02, 95% CI: 1.02, 1.03), CVD (HR: 1.03, 95% CI: 1.03, 1.04), dementia (HR: 1.04, 95% CI: 1.03, 1.05), liver disease (HR: 1.06, 95% CI: 1.05, 1.07), respiratory disease (HR: 1.07, 95% CI: 1.06, 1.07), chronic kidney disease (HR: 1.09, 95% CI: 1.09, 1.10), hypertension (HR: 1.05, 95% CI: 1.04, 1.06), type 2 diabetes mellitus (HR: 1.07, 95% CI: 1.06, 1.08), eyes disease (HR: 1.02, 95% CI: 1.01, 1.02), depression (HR: 1.03, 95% CI: 1.02, 1.03), and anxiety (HR: 1.02, 95% CI: 1.01, 1.02), with an exception of osteoarthritis (HR: 1.00, 95% CI: 1.00, 1.01) (Supplementary Table 9). The Kaplan-Meier survival curves exhibited discernible trajectories among the quartile groups of MetaboAgeMortAccel (Fig. 3b and Supplementary Fig. 3b). The group in the highest quartile (Q4) showed a significant association with increased risks of multiple disease incidences compared to the lowest quartile (Q1) (Supplementary Table 10 and 11).

### Discriminative improvements beyond clinical predictors

Based on the findings depicted in Fig. 2d, it is evident that the area under the curve (AUC) of MetaboAgeMort displayed a considerable improvement relative to chronological age, aligning it more closely with the AUC of the conventional risk factors model. MetaboAgeMort added predictive utility of 10-year mortality beyond conventional risk factors (i.e., chronological age, sex, alcohol intake frequency, smoking status, BMI, systolic blood pressure, triglycerides, creatinine, total cholesterol, HDL cholesterol, and prevalent diabetes, CVD and cancer)^11^. Compared with the conventional risk factors model, the combined model including MetaboAgeMort had better discrimination ability, as demonstrated by significantly increased C-statistics (0.017, *P* < 0.001) (Supplementary Table 12). The superior performance of MetaboAgeMort was further confirmed through substantial enhancements in reclassification, as evaluated by integrated discrimination improvement (IDI: 0.018, 95% CI: 0.004, 0.021) (Supplementary Table 12), suggesting that MetaboAgeMort captures something above and beyond what can be explained for mortality risk by conventional risk factors.

### The comparison of MetaboAgeMort with MetaboAge and MetaboHealth score

In addition, we calculated two pre-existing well-known multi-metabolite scores (i.e., MetaboAge^5^ and MetaboHealth score^12^), and assessed their correlations with the mortality risk and multiple diseases incidence. The MetaboAge score predicts chronological age (in years) directly. Two versions of MetaboAge were calculated: MetaboAge_LM (generated through linear regression), and MetaboAge_EN (generated through ElasticNET regression)^20^. The MetaboHealth score is a multivariate model predicting all-cause mortality. It was calculated as the weighted sum of 14 log-transformed and cohort-scaled metabolites. The distributions of these multi-metabolite scores in the testing set are shown in Supplementary Fig. 5a. As shown in Supplementary Table 13, after adjustment for potential confounders, the highest quartile group (Q4) of the MetaboHealth score has significantly increased risks of all-cause mortality, cause-specific mortality, and multiple incident diseases, compared to the lowest quartile group (Q1). The MetaboAge_EN showed significant association with all-cause mortality, CVD mortality, and several cardiometabolic diseases (i.e., CVD and T2DM). Conversely, MetaboAge_LM did not demonstrate any appreciable relationship with these same clinical endpoints. As shown in Supplementary Table 14, even after mutual adjustment, both MetaboAgeMort and MetaboHealth score maintained significant correlations with all-cause mortality. Notably, the Akaike Information Criterion (AIC) for the model integrating MetaboAgeMort indicated a relatively superior fit, implying that MetaboAgeMort may have a higher predictive capacity or enhanced explanatory power in forecasting all-cause mortality when compared to the model integrating the MetaboHealth score.

Moreover, we compared MetaboAgeMort with these multi-metabolite scores, in terms of their predictive utility for 10-year mortality risk and multiple diseases incidence. As shown in Supplementary Fig.5b, MetaboAgeMort displayed a significantly higher AUC than that of the MetaboAge and MetaboHealth score for 10-year all-cause mortality prediction (*P* < 0.001). Combining MetaboAgeMort with conventional risk factors resulted in an AUC that outperformed models integrating MetaboAge (MetaboAge_EN and MetaboAge_LM, both P < 0.001) or MetaboHealth score (P = 0.021) with the same conventional risk factors. When combining MetaboAgeMort with MetaboHealth score and conventional risk factors, the resulting AUC was higher than that of the model integrating only MetaboHealth score with conventional risk factors, but this difference was not significant when compared to the model integrating only MetaboAgeMort with conventional risk factors. This finding suggests that MetaboAgeMort may capture something beyond what can be explained for mortality risk by MetaboHealth score, when taking into account conventional risk factors. MetaboAgeMort also exhibited a higher predictive utility for multiple diseases (except for cancer incidence) compared to the MetaboHealth score (Supplementary Fig. 5c). After accounting for chronological age and sex, the AUC of the model integrating both MetaboAgeMort and MetaboHealth score was higher than that of the model with MetaboHealth score alone, but did not show a significant improvement over the model with MetaboAgeMort alone. This finding suggests that MetaboAgeMort may also capture something beyond what can be explained for diseases incidence by MetaboHealth score.

### Modifiable factors for MetaboAgeMort

Then we investigated the modifiable factors of MetaboAgeMort. We considered a total of 107 potentially modifiable factors from the UKB baseline survey. After Bonferroni correction, 99 factors showed a significant association with MetaboAgeMort (*P* < 4.67×10^-4^, Fig. 4a and Supplementary Table 13). For 16 factors across four categories, the associations were relatively more substantial (≥ 2 years of change in MetaboAgeMort) per 1-SD change in the factor (Supplementary Fig. 6), such as forced vital capacity (FVC) (tertile 3 vs. tertile 1, β =[-2.22, 95% CI:[−2.28, −2.15), body fat percentage (tertile 3 vs. tertile 1, β =[3.55, 95% CI: 3.49, 3.61), dietary index (tertile 3 vs. tertile 1, β =[−2.04, 95% CI: −2.09, −1.98), and average total household income before tax (greater than 100,000 vs. less than 18,000, β =[−3.34, 95% CI: − 3.44, −3.24). Comparable correlations were noted when stratifying by chronological age and sex (Supplementary Table 13).

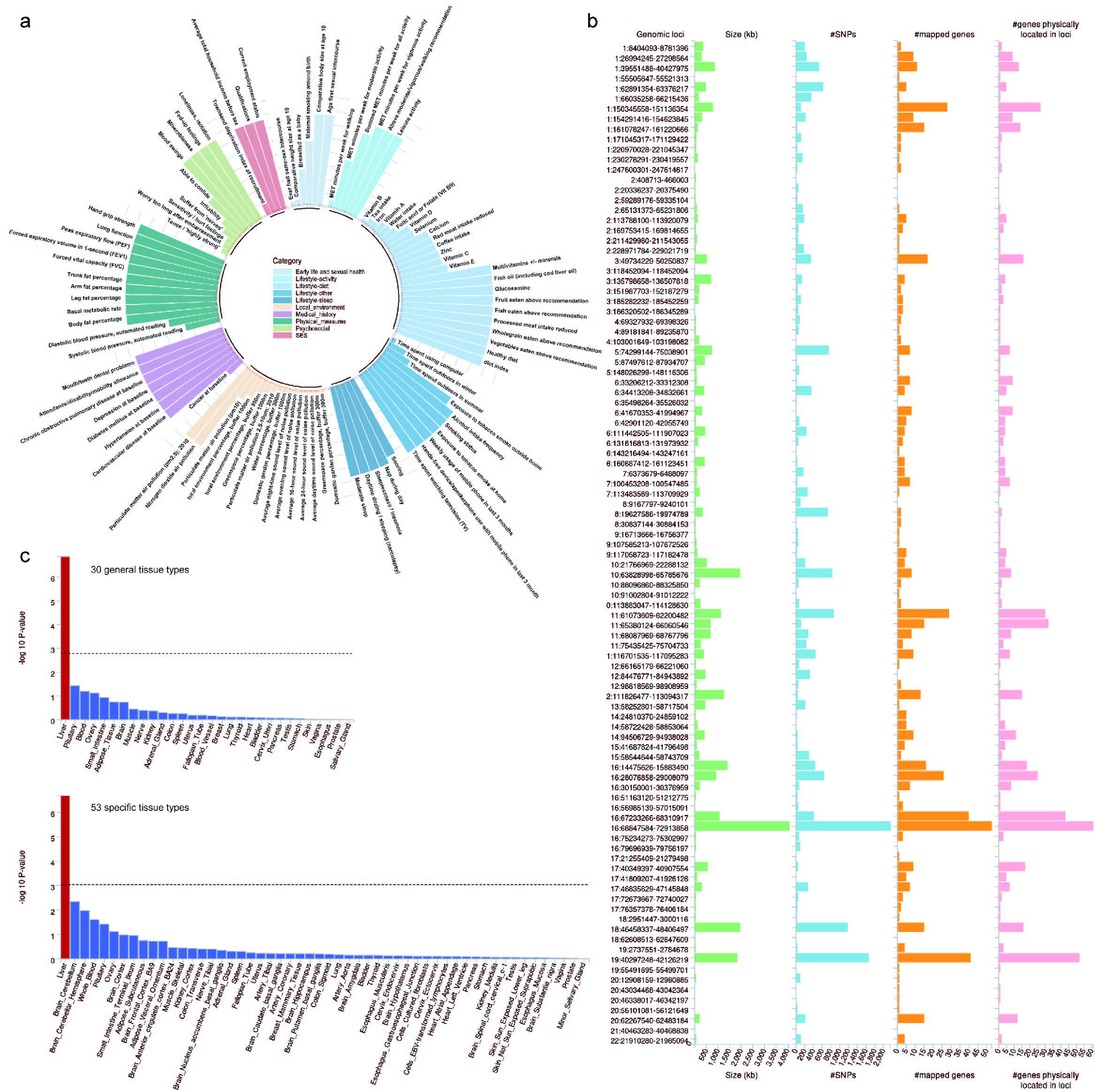

### Genetic determinants for MetaboAgeMort

To better understand the genetic mechanisms underlying metabolomic aging, we performed a genome-wide association study (GWAS) analysis of MetaboAgeMort. In the GWAS analysis, 11,688 SNPs significantly associated with MetaboAgeMort were identified (*P* < 5 × 10^-8^) (Supplementary Fig. 7). The SNP-derived heritability of MetaboAgeMort was 38.26% (*P* = 2.23 × 10^-86^). Using the Functional Mapping and Annotation (FUMA) online platform (v1.5.2), we pinpointed 1,068 independent significant SNPs, 319 lead SNPs, and 99 genomic risk loci (Supplementary Table 14-17). Furthermore, 585 prioritized genes that may be involved in the genetic etiology of MetaboAgeMort were identified by positional mapping (Supplementary Table 18). The leading SNP of the most significant locus (rs174575, locus 57) was positioned near or within *FADS1* and *FADS2* on chromosome 11. The leading SNP of the second most significant locus (rs217184, locus 78) was in *TXNL4B*, *HPR*, and *HP* on chromosome 16 (Supplementary Table 19). Summary results per genomic risk locus are shown in Fig. 4b.

We also performed analyses pertaining to genes and tissue enrichment using Multi-Marker Analysis of GenoMic Annotation (MAGMA) v1.08 within FUMA. In genome-wide gene-based association analysis (GWGAS), 310 genes were determined to be genome-wide significant after applying Bonferroni correction (Supplementary Table 20), where 271 genes were also positionally mapped to significant loci from the SNP-based analysis above. The gene-set analysis identified 13 significant gene sets after Bonferroni correction, including lipid-related biological process (e.g., reverse cholesterol transport, cholesterol metabolism, and phospholipid homeostasis), CYP2E1 reactions, and liver specific genes (Supplementary Table 21). The tissue-enrichment analysis indicated that liver displayed significant specificity in gene expression for the MetaboAgeMort-associated genes (Fig. 4c and Supplementary Table 22-23).

## Discussion

Leveraging the large-scale metabolomics data, we have formulated a groundbreaking aging measure, MetaboAgeMort, based on 10-year all-cause mortality risk prediction. The MetaboAgeMort demonstrated remarkable predictive utility of mortality risk across a wide range of demographic and socioeconomic stratifications, as well as health behavior factors and causes of death. Significantly, the conventional risk factors were augmented by MetaboAgeMort in terms of predictive accuracy for 10-year mortality. Meanwhile, our study has revealed compelling associations between accelerated metabolomic aging within the same age group and an increased likelihood of various health-related outcomes. This implies that it could serve as a comprehensive indicator for health and mortality risk stratification in a clinical setting. Next, we identified 99 modifiable factors across seven categories for MetaboAgeMort, highlighting the crucial role played by body composition, healthy diet, SES, and pulmonary function in the process of metabolomic aging. The genetic analyses ultimately revealed 99 significant genomic risk loci and 271 genes linked to MetaboAgeMort, thus offering new insights into the genetic architecture of metabolomic aging. Previous studies have generated metabolomic clocks while using chronological age as a surrogate^21^.

In comparison, our MetaboAgeMort further incorporates information on all-cause mortality risk, which is considered a more reliable surrogate for biological aging than chronological age, through a sophisticated modeling method. As one would expect from a measure of aging, MetaboAgeMort not only assesses the risks of all-cause and cause-specific mortality, but also the risks of various incident diseases, highlighting its considerable potential in the early detection of individuals at risk and facilitating timely and effective interventions. It is worth mentioning that we have substantiated the contribution of MetaboAgeMort in enhancing the predictive capacity for 10-year all-cause mortality risk in addition to conventional risk factors. These findings translate into potential clinical application of MetaboAgeMort as an additional source of discriminatory information to refine comprehensive risk assessments for death and diseases. To date, numerous studies have endeavored to identify metabolite predictors of mortality risk and have successfully developed multivariate metabolite scores (e.g., MetaboHealth score) that exhibited significant association with all-cause mortality^22^. Nonetheless, the utilization of these metabolite scores is constrained due to the reliance on cohort-specific scaled biomarker values, thereby impeding its clinical application in identifying individuals with a propensity for accelerated aging and comparison across different populations. MetaboAgeMort, directly calculated from individual-level data, more intuitively shows the heterogeneity in mortality risk among individuals of the same chronological age, exhibiting enhanced generalization.

The 35 metabolic biomarkers employed in our study to develop MetaboAgeMort are implicated in diverse processes, including lipoprotein and fatty acid metabolism, fluid balance, and inflammation, indicating that aging is an intricate multidimensional phenomenon. Previous studies have sought to elucidate the interaction between metabolites and aging. In line with the discoveries made by Deelen et al.^12^, our study revealed that average diameter for VLDL particles, histidine, leucine, phenylalanine, valine, glucose, acetoacetate, albumin, glycoprotein acetyls, the ratio of polyunsaturated fatty acids to total fatty acids (i.e., omega-3 fatty acids to total fatty acids percentage, and linoleic acid to total fatty acids percentage), and total lipids in small HDL were essential independent indicators for mortality. In addition, we observed associations for several other metabolic biomarkers, such as ketone bodies and relative lipoprotein lipid concentrations. KBs are endogenous fuels generated by the liver in response to metabolic stress^23^. In a healthy community-based population, higher elevated endogenous KBs have shown to be positively associated with all-cause mortality^24^. Relative lipoprotein lipid concentrations play a role in lipid homeostasis and their associations with mortality may be partially attributed to their regulatory effect on plasma triglyceride levels, a critical mortality risk factor^25^. Collectively, our study contributes to the comprehension of metabolic alterations that underlie the process of aging.

The primary focus of aging research has been on the development of strategies to combat aging. Measures like MetaboAgeMort, which capture future morbidity and mortality risk, could facilitate evaluation of intervention efficacy while eliminating the requirement for extended follow-up periods. A recent study in the UK Airwave cohort has discovered a correlation between metabolomic aging and several factors such as overweight, obesity, heavy drinking, diabetes, depressive symptoms, depression, anxiety, and post-traumatic stress disorder^7^. Using a larger-scale population-based cohort, our study meticulously investigated the modifiable factors associated with metabolomic aging. Consequently, we identified a total of 99 potential factors across seven distinct categories: SES, early life and sexual health, medical history, physical measures, psychosocial factors, local environment, and lifestyle. Stronger associations (β > 2) were quantitatively observed for 16 factors related to pulmonary function, body composition, SES, dietary quality, smoking status, alcohol intake, and disease status, all of which have previously been reported to be associated with aging^26–29^. Our study provides a metabolomic insight into the mechanisms linking these factors to aging process. Certain local environmental factors, such as fine particulate matter (PM2.5) and nitrogen dioxide (NO2), have been evidenced to potentiate oxidative stress responses within biological systems, impair mitochondrial function, and subsequently lead to metabolic irregularities^30,31^. The correlation between these factors and MetaboAgeMort underscores the significance of enacting urban planning and environmental preservation policies to slow down the aging process. Furthermore, MetaboAgeMort demonstrated responsiveness to specific early-life exposure factors, suggesting that targeted interventions during the critical developmental period, such as optimizing maternal nutrition and improving the developmental environment for children, could potentially contribute to the promotion of healthy aging and reduction of risks associated with late-life mortality and morbidity^32^.

Genetics play a substantial role in determining individual biological aging rates^33^. Based on our current understanding, this study offers the initial evidence regarding the genetic determinants of metabolomic aging. By utilizing genotyping data, we pinpointed 99 genomic risk loci and 271 genes associated with MetaboAgeMort. The most significant SNPs were identified within the *FADS* cluster (*FADS1*, *FADS2*) on chromosome 11. These genes have emerged as significant genes in prior studies on serum omega-3 fatty acid^34^, a crucial type of polyunsaturated fatty acids that have demonstrated favorable impacts on age-related diseases (e.g., CVD and metabolic diseases)^35^. We also identified several genome-wide significant genes on chromosome 16, such as *TXNL4B*, *HPR*, and *HP*. These genes play integral roles in modulating the core biological mechanisms, including cell cycle progression, oxidative balance maintenance, protein conformational dynamics and stability, which all exhibit profound interdependencies with the aging process^36,37^.Moreover, the MAGMA gene-set analysis unveiled the critical involvement of lipid metabolism, CYP2E1-mediated reactions, and liver-specific genes in metabolomic aging, providing valuable insights into the underlying molecular mechanisms and potential therapeutic targets pertinent to this process. The tissue-enrichment analysis further emphasizes the importance of liver in metabolomic aging. This finding implies that strategies focused on preserving or restoring liver health, such as modulating key metabolic pathways, enhancing antioxidant defenses, and stem cell therapy, may have far-reaching systemic benefits in countering metabolomic aging process. Further research is needed to refine these potential strategies and evaluate their efficacy in promoting healthy aging and preventing age-related diseases. Notably, the pathways identified for MetaboAgeMort were distinct from those enriched by genes associated with PhenoAgeAccel or BioAgeAccel^16^, thus reaffirming that the heterogeneous aging patterns observed among individuals might be partly attributed to varying genetic susceptibilities.

Some limitations in this study should also be noted. First, the number of biomarkers captured by the targeted NMR platform is only a fraction of the metabolites in the human plasma. Nevertheless, NMR has the ability to offer highly accurate quantification at a minimal expense, thus facilitating the straightforward implementation of metabolomic clocks in population health. Second, the association between modifiable risk factors and MetaboAgeMort is cross-sectional, and further causal inferences are needed. Third, even though we have taken into account numerous modifiable risk factors, it is conceivable that certain factors may have been unintentionally neglected. Fouth, the majority of participants in the UKB were White British and tended to be healthier and wealthier^38^, and thus, our sample was less representative of the overall UK adult population. It is worth mentioning that the generalizability of our findings to populations in developing countries or other contexts may be limited.

In conclusion, we have successfully developed a novel metabolomic-based measure of aging known as MetaboAgeMort. This measure has proven to be highly predictive of mortality and various diseases. Of particular significance is the fact that MetaboAgeMort can enhance the predictive accuracy of 10-year mortality beyond conventional risk factors. These findings imply that it has remarkable potential as a comprehensive measure of overall health and risk of mortality in a clinical setting. The potential of body composition, healthy diet, SES, pulmonary function, smoking status, and alcohol intake were highlighted as possible factors for delaying metabolomic aging. Our research has led us to the identification of 99 significant genomic risk loci and 271 genes linked to MetaboAgeMort. This breakthrough sheds new light on the complex genetic underpinnings governing metabolomic aging processes.

## Methods

### Study participants

The UKB is a large prospective cohort study that comprised over 500,000 participants aged 37-73 years at the time of baseline assessment (2006-2010). Information was collected via touch-screen questionnaires, biological samples, physical measurements, and linked medical or death register records. Detailed study design and methodology were described elsewhere^39^. Ethics for the UKB was approved by the North West Multicenter Research Ethics Committee, and all participants have provided signed informed consent.

### Plasma metabolomics

A total of 251 metabolic biomarkers for EDTA plasma samples from a randomly selected subset of approximately 280,000 UKB participants were measured between June 2019 and April 2020 (Phase 1) and April 2020 and June 2022 (Phase 2) using a high-throughput NMR metabolomics platform developed by Nightingale Health Ltd. The metabolic biomarkers span multiple metabolic pathways, including fatty acids, fatty acid compositions, and lipoprotein lipids in 14 subclasses, as well as various low-molecular weight metabolites, such as ketone bodies, amino acids, and glycolysis metabolites. Detailed protocols for sample collection and methodology for the Nightingale NMR pipeline were described elsewhere^40,41^. The present study considered 249 available metabolic biomarkers (except for glucose-lactate and spectrometer-corrected alanine) and the values of each metabolic biomarker were transformed using natural logarithmic transformation (ln[x+1]) followed by Z-normalisation prior to analysis.

### Development of MetaboAgeMort and MetaboAgeMortAccel

Two sequential steps were undertaken in order to create the MetaboAgeMort model. In the first step, we identified aging-related metabolic biomarkers. Initially, We evaluated the associations of each metabolic biomarker (per 1-SD increment) with all-cause mortality using multivariable Cox regression models, with adjustment for chronological age, sex, ethnicity, education level, Townsend deprivation index (TDI), alcohol intake frequency, smoking status, regular exercise, healthy diet, BMI, cholesterol-lowering medication, anti-hypertensive medication, anti-diabetes medication, and prevalent diseases at baseline (i.e., cancer, CVD, hypertension, diabetes mellitus, and chronic obstructive pulmonary disease [COPD]) among all participants (n = 239,291), and 185 metabolic biomarkers were selected with Bonferroni correction. Then, we employed a Cox regression model with LASSO penalization--where the hazard of all-cause mortality was regressed on 185 metabolic biomarkers and chronological age--in the training set. Finally, an optimal λ of 0.00097 were selected via ten-fold cross-validation, and 36 variables, including chronological age were assigned nonzero coefficients.

In the second step, we constructed the MetaboAgeMort in the training set by adopting the methodology previously proposed by Levine et al.^18^. We fitted two proportional hazards regression models based on the parametric Gompertz distribution: one used 36 variables selected above as predictors, and the other used only chronological age as a predictor. Based on the two models, we predicted the 10-year all-cause mortality risk using the cumulative distribution function, respectively. We then converted the mortality risk into units of years by equating the risk from the two models and solving for age, thus obtaining MetaboAgeMort. In general, an individual’s MetaboAgeMort represents the chronological age within the general population corresponding to that individual’s mortality risk. For example, two individuals are chronologically 40 years old, but one may have a MetaboAgeMort of 45 years and the other a MetaboAgeMort of 35 years, indicating that they have the average mortality risk of someone who is chronologically 45 or 35 years old, respectively. In addition, we calculated a metric, MetaboAgeMortAccel, following the methods by Liu et al^42^. MetaboAgeMortAccel represents the divergence of MetaboAgeMort from chronological age, defined as the residual resulting from a least-squares linear model when regressing MetaboAgeMort on chronological age.

### Health-related outcomes

Information on date and cause of death were obtained through the linkage to national death registries. The all-cause and cause-specific mortality (i.e., cancer, CVD, respiratory disease, neurodegenerative disease, digestive disease, and other causes) were determined using the International Classification of Disease (ICD)-10 codes (Supplementary Table 24)^43^. Follow-up time was calculated from the date of baseline assessment to the date of death, loss to follow-up, or end of follow-up (Dec 31, 2022), whichever came first.

Information on diagnoses and medical conditions of the participants were obtained through the linked hospital inpatient record data, self-reported data, and primary care data from the UK National Health Services. The incident diseases were ascertained by the ICD-9 and ICD-10 codes (Supplementary Table 25). Follow-up time was calculated from the date of baseline assessment to the date of first diagnosis of the disease, death, loss to follow-up, or end of follow-up (Oct 31, 2022), whichever came first.

### MetaboAge and MetaboHealth score

The MetaboAge score predicts CA (in years) directly^5^. Two different models were utilized: a linear regression model (MetaboAge_LM) and an ElasticNET regression model (MetaboAge_EN). The model weights/coefficients were obtained from the most recent publication by Akker et al.^20^ In quality control, we excluded three samples with one or more zero values per sample, as well as one or more concentrations that were more than 5 times the SD away from the overall mean of the feature. The MetaboHealth score is a multivariate model predicting all-cause mortality^12^. It was calculated as the weighted sum of 14 log-transformed and cohort-scaled metabolites, using the R-package MiMIR^44^. To avoid infinite values after log-transformation, a value of 1was added to the metabolites containing any zero.

### Modifiable factors

We have taken into account a total of 107 potentially modifiable factors from the UKB baseline survey. Details of processing the factors are presented in Supplementary Table 26. These factors were classified into seven categories: local environment (e.g., greenspace percentage, buffer 1000m), psychosocial (e.g., nervous feelings), SES (e.g., TDI), medical history (e.g., prevalent CVD at baseline), early life and sexual health (e.g., breastfed as a baby), physical measures (e.g., handgrip strength), and lifestyle (e.g., healthy diet).

### Genome-wide association analysis

To better understand the genetic mechanisms underlying MetaboAgeMort, we performed a GWAS analysis using the data from UKB v3 genotyping release. The SNPs were excluded if meeting any of the following criteria: (1) minor allele frequency < 0.01, (2) Hardy-Weinberg equilibrium test *P* < 1.0×10^-6^, (3) missing minor allele frequency, or results of Hardy-Weinberg equilibrium test. This particular aspect of the analysis was constrained to individuals of White British descent (n= 226,937). The fastGWA-MLM analysis in the Genome-wide complex trait analysis (GCTA) software (version 1.94.1) was used to perform GWAS analysis^45^. Models included chronological age, sex, genotype array, and the top 10 principal components as the covariates. The genome-based restricted maximum likelihood (GREML) method in GCTA was used to estimate the SNP-based heritability (variance explained by all the SNPs)^46^.

### Functional mapping and annotation

The FUMA online platform (v1.5.2) was applied for functional mapping and annotation of GWAS results (default parameters were used unless explicitly stated otherwise), with annotations derived from the human genome assembly GRCh37 (hg19)^47^. To identify independent genomic risk loci (defined by r^2^ > 0.6) and variants in linkage disequilibrium (LD) with lead SNPs, the SNP2Gene module was applied using the genetic data of European populations in 1000G phase3 as the reference^48^. Positional mapping with a 10 kilobase (kb) window size was employed to map risk loci to neighboring protein-coding genes.

### Gene-based association, gene-set, and gene-property analyses with MAGMA

We performed analyses pertaining to genes and tissue enrichment (i.e., GWGAS, gene-set, and tissue expression analyses) using MAGMA v1.08 within FUMA^49^. For each of the 18,955 protein-coding genes, GWGAS accessed the joint effect of all variants within the gene. Bonferroni correction was used to establish the genome-wide significance threshold (*P* < 0.05/ 18,955 = 2.64×10^-6^). Gene-set analysis was further performed using hypergeometric tests for curated gene sets and Gene Ontology (GO) terms from MsigDB v7.0 to discern whether specific biological pathways or cellular functions were implicated in the genetic etiology of MetaboAgeMort, with Bonferroni correction also being utilized. Simultaneously, tissue enrichment analysis was carried out with 30 general and 53 specific tissue types from GTEx v8^50^.

### Covariates

Information on chronological age, sex (female or male), ethnicity (White or Non-white), education level (high, intermediate, or low), alcohol intake frequency (never or special occasions only, one to three times per month, one to four times per week, or daily or almost daily), smoking status (never, previous, or current smoker), regular exercise (yes or no), healthy diet (yes or no), and medication information were collected through questionnaire interview. The TDI was assigned by participants’ postcodes, representing SES levels^51^. BMI (kg/m^2^) was calculated as measured weight/height^2^.

### Statistical analysis

Baseline characteristics were described using median (interquartile range, IQR) or count (percentage). Two-sample Wilcoxon test for continuous variables and chi-squared test for categorical variables were used to test the differences between the training and the testing set.

The study’s roadmap is illustrated in Fig. 1. The Cox proportional hazard model was used to evaluate the association between MetaboAgeMort and all-cause mortality, with adjustment for chronological age, sex, ethnicity, education level, TDI, alcohol intake frequency, smoking status, regular exercise, healthy diet, BMI, medication use, and prevalent diseases at baseline. To further assess robustness, we repeated the analyses by (1) stratified by several demographic, socioeconomic, as well as health behavior factors, (2) excluding participants who died within 5 years of follow-up to reduce the influence of end-of-life metabolomic status, and (3) only including participants who were free of prevalent diseases at baseline to minimize the influence of reverse causality. Proportional hazards of the associations were tested using Schoenfeld’s residuals. We then grouped the participants into quartiles of MetaboAgeMortAccel. The Kaplan-Meier plots were drawn to visualise survival curves.

The evaluation of MetaboAgeMort associations with cause-specific mortality and incident diseases was conducted using Fine and Gray’s competing risk models^52^. When constructing models evaluating the associations of MetaboAgeMort with incident diseases, we excluded the participants with a specific diagnosis before or at the time of recruitment from the models. Chronological age, sex, ethnicity, education level, TDI, alcohol intake frequency, smoking status, regular exercise, healthy diet, and BMI were used as covariates. The associations of MetaboAgeMortAccel with mortality and incident diseases were evaluated using same method as that for MetaboAgeMort. The second and the third MetaboAgeMortAccel quartiles (Q2 and Q3) were set as the reference.

Next, receiver operating characteristic (ROC) curves were used to evaluate the utility of MetaboAgeMort for 10-year all-cause mortality risk prediction beyond conventional risk factors. C-statistic and IDI were calculated, in comparison to that of the conventional risk factors model. In addition, we used ROC to compare its predictive utility for 10-year all-cause mortality risk and multiple diseases incidence with several previously trained multi-metabolite scores (i.e., MetaboAge and MetaboHealth score). Moverover, we tested the associations of these multi-metabolite scores with cause-specific mortality and diverse age-related diseases using the same methods employed for MetaboAgeMort.

Multivariable linear regression models were applied to test the response of MetaboAgeMort (response variable) for each modifiable factor (independent variable), with a Bonferroni-corrected significance threshold for identifying top hits (*P* < 0.05/107 = 4.67×10^-4^). In these analyses, continuous variables were Z-normalised, and results were shown as β coefficients per 1-SD increment in the corresponding factor (based on the availability of each individual factor). Moreover, we divided modifiable factors into tertiles and showed the results using the lowest tertile as a reference. The models were adjusted for chronological age, sex, and ethnicity. Stratified analyses according to chronological age (< 60 and ≥ 60 years) and sex (male and female) were conducted, utilising Bonferroni-correction method to determine top hits. Data analyses and visualisation were all performed in R version 4.2.2. A two-sided *P* of ≤ 0.05 was considered statistically significant.

## Funding

This research was supported by Grants from Research Center of Prevention and Treatment of Senescence Syndrome, School of Medicine Zhejiang University (2022010002), “Pioneer” and “Leading Goose” R&D Programs of Zhejiang Province (2023C03163), the National Natural Science Foundation of China (72374180), the Fundamental Research Funds for the Central Universities, Key Laboratory of Intelligent Preventive Medicine of Zhejiang Province (2020E10004), and Zhejiang University Global Partnership Fund (188170-11103). The funders had no role in the study design; data collection, analysis, or interpretation; in the writing of the report; or in the decision to submit the article for publication.

## Data Availability

All data produced are available online at www.ukbiobank.ac.uk/register-apply.

## Acknowledgments

This research was conducted using the UK Biobank resource under application number 61856. We wish to acknowledge the UK Biobank participants who provided the sample that made the data available.

## Competing interests

The authors have no conflicts of interest related to this manuscript to declare.

## Ethics approval and consent to participate

UK Biobank has approval from the North West Multi-Centre Research Ethics Committee as a Research Tissue Bank approval in 2011 and is renewed every 5 years, which allowed researchers to use data from UK Biobank without an additional ethical clearance. All participants have provided signed informed consent.

## Notes

### Competing Interest Statement

The authors have declared no competing interest.

### Funding Statement

This study was funded by Grants from Research Center of Prevention and Treatment of Senescence Syndrome, School of Medicine Zhejiang University (2022010002), "Pioneer" and "Leading Goose" R&D Programs of Zhejiang Province (2023C03163), the National Natural Science Foundation of China (72374180), the Fundamental Research Funds for the Central Universities, Key Laboratory of Intelligent Preventive Medicine of Zhejiang Province (2020E10004), and Zhejiang University Global Partnership Fund (188170-11103). The funders had no role in the study design; data collection, analysis, or interpretation; in the writing of the report; or in the decision to submit the article for publication.

### Author Declarations

This research was conducted using the UK Biobank resource under application number 61856. The North West Multi-Centre Research Ethics Committee gave ethical approval for the UK Biobank and is renewed every 5 years, which allowed researchers to use data from UK Biobank without an additional ethical clearance. All participants have provided signed informed consent.

